# Development of a machine-learning model for therapeutic efficacy prediction of preoperative treatment for esophageal cancer using single nucleotide variants of autophagy-related genes

**DOI:** 10.1101/2024.09.07.24313244

**Authors:** Yutaka Miyawaki, Masataka Hirasaki, Yasuo Kamakura, Tomonori Kawasaki, Yasutaka Baba, Tetsuya Sato, Satoshi Yamasaki, Hisayo Fukushima, Kousuke Uranishi, Yoshinori Makino, Hiroshi Sato, Tetsuya Hamaguchi

**Affiliations:** Department of Gastroenterological Surgery, Saitama Medical University International Medical Center, 1397-1 Yamane, Hidaka, Saitama, 350-1298, Japan; Department of Clinical Cancer Genomics, Saitama Medical University International Medical Center, 1397-1 Yamane, Hidaka, Saitama 350-1298, Japan; Division of Biomedical Sciences, Research Center for Genomic Medicine, Saitama Medical University, 1397-1 Yamane, Hidaka, Saitama 350-1298, Japan; Department of Pathology, Saitama Medical University International Medical Center, 1397-1 Yamane, Hidaka, Saitama 350-1298, Japan; Department of Diagnostic Radiology, Saitama Medical University International Medical Center, 1397-1 Yamane, Hidaka, Saitama 350-1298, Japan; Biomedical Research Center, Faculty of Medicine, Saitama Medical University, 1397-1 Yamane, Hidaka, Saitama 350-1298, Japan; Department of Medical Oncology, Gastroenterological Oncology, Saitama Medical University International Medical Center, 1397-1 Yamane, Hidaka, Saitama 350-1298, Japan

**Keywords:** Biomarker, Esophageal Cancer, Machine Learning, Neoadjuvant Therapy, Recurrence

## Abstract

Neoadjuvant chemotherapy with cisplatin + 5-fluorouracil followed by radical surgery is the standard treatment for stage II and III esophageal cancers. Although, a more potent regimen comprising cisplatin + 5-fluorouracil with docetaxel, has shown superiority in overall survival compared to the cisplatin + 5-fluorouracil regimen, it involves worsening of Grade 3 or higher adverse events due to docetaxel. Based on these reports, this study aimed to construct a prognostic system for cisplatin + 5-fluorouracil regimens, particularly for locally advanced cancers, to guide selection of neoadjuvant chemotherapy. Biopsy specimens from 82 patients who underwent a cisplatin + 5-fluorouracil regimen plus radical surgery at Saitama Medical University International Medical Center between May 2012 and June 2020 were analyzed. Variants in 56 autophagy- and esophageal cancer-related genes were identified using targeted enrichment sequencing. Overall, 13 single nucleotide variants, including eight non-synonymous group single nucleotide variants predicting recurrence were identified using Fisher’s exact test with recurrence as a two-group event, which showed a significant difference (p < 0.05). Additionally, machine learning was used to predict recurrence using 21 features, including eight patient backgrounds. The results showed that the Naive Bayes was highly reliable with an accuracy of 0.88 and Area Under the Curve of 0.9. Thus, we constructed a machine learning model to predict recurrence in patients with esophageal cancer treated with a cisplatin + 5-fluorouracil regimen. We believe that our results will provide useful guidance for the selection of neoadjuvant adjuvant chemotherapy, including the avoidance of docetaxel.

## Introduction

Esophageal cancer ranks seventh in terms of incidence and sixth in terms of overall mortality among all cancers^1^. The conventional standard treatment for stages II and III esophageal cancer in Japan is neoadjuvant chemotherapy with cisplatin + 5-fluorouracil (CF), followed by radical surgery^2^. According to previous reports, the 5-year survival rate for stage II cancers after neoadjuvant chemotherapy with the CF regimen is 69%. However, the 5-year survival rate for stage III cancer is poor at 52%, indicating that neoadjuvant chemotherapy with CF regimen has a limited effect in locally advanced cases^3,4^. Therefore, a more potent neoadjuvant chemotherapy regimen with docetaxel + CF (DCF) has attracted attention in recent years. A phase III trial (JCOG1109) comparing the superiority of neoadjuvant chemotherapy with DCF and CF regimens demonstrated an overall survival (OS) advantage in the neoadjuvant DCF arm^5^. Based on these results, neoadjuvant DCF therapy became the standard treatment in Japan in February 2022^6^. However, the exacerbation of adverse events with docetaxel, specifically grade 3 or higher leukopenia (6.7–63.8%), neutropenia (23.4–85.2%), and hyponatremia (6.2– 26.0%), have also been reported simultaneously^5^. The high level of chemotherapy-related adverse events may potentially force a series of treatment interruptions, prevent the maintenance of ideal chemotherapy dose intensity, and make it difficult to complete treatment, including subsequent surgery. Chemotherapy-induced leukopenia (CIL) is also a known prognostic factor of chemotherapy in some malignancies, although it is not currently evident in esophageal cancer ^7–9^. Based on these results, we believe that the establishment of prognostic markers, especially for CF regimens for cT3 resectable advanced esophageal cancers, will provide useful guidance in the selection of neoadjuvant chemotherapy, including the avoidance of docetaxel administration.

Autophagy is a highly regulated process of degradation and recycling of cellular components. The most important feature of autophagy is that it degrades intracellular proteins and organelles and recycles them as a new source of nutrients^10^. Recently, autophagy was shown to contribute to the acquisition of chemotherapy resistance in established cancers via intracellular recycling, provide a substrate for metabolism, and maintain a functional pool of mitochondria^11^. In patients with esophageal cancer receiving CF and DCF regimens, a high expression of PINK1, an initiator of mitophagy, was associated with poor prognosis, suggesting that PINK1-mediated mitophagy contributed to resistance to neoadjuvant therapy^12^. However, it was not established as a biomarker because high PINK1 protein expression did not correlate with the response to neoadjuvant chemotherapy in biopsy specimens obtained before neoadjuvant chemotherapy. In contrast, we previously reported that single-nucleotide variants in PINK1 may be biomarkers for non-recurrence in patients with colorectal cancer treated with postoperative adjuvant chemotherapy^13^.

This study aimed to construct a prognostic system for CF regimens, particularly for locally advanced cancers. The single nucleotide variants (SNVs) and insertions and deletions (INDELs) for autophagy- and esophageal cancer-related genes were identified in a sample of patients who received neoadjuvant chemotherapy with a CF regimen for esophageal squamous cell carcinoma (ESCC). The identified SNVs and INDELs were statistically examined, with recurrence as an event between the two groups. Finally, a machine-learning model was constructed to predict recurrence by comprehensively considering 13 variants that were significantly correlated with recurrence. This provided a useful guidance for the selection of neoadjuvant chemotherapy, including avoiding docetaxel administration.

## Material and methods

### Tissue samples

Ninety-one patients with esophageal cancer who underwent a neoadjuvant CF regimen plus radical surgery at Saitama Medical University International Medical Center between May 2012 and June 2020 were eligible. Of these, samples from 82 patients with sufficient DNA content were included in this study　(Table 1). The location of tumor cells in the tissue specimen was determined both visually and microscopically by a pathologist using hematoxylin and eosin-stained (H&E) sections, which were taken from paraffin blocks of biopsy specimens of the esophageal cancer tissue.

**Table 1.** Clinical characteristics of the patients included in this study.

| **n=82** |  |
| --- | --- |
| **Age years** |  |
| Median (range) | 68 (51-80) |
| **Gender (%)** |  |
| Male / Female | 74 (90%) / 8 (10%) |
| **Organization type (%)** |  |
| Basaloid/SCC | 3 (4%) / 79 (96%) |
| **Neo-adjuvant course (%)** |  |
| 1 / 2 | 11 (13%) / 71 (87%) |
| **Tumor location (%)** |  |
| Upper / Middle / Lower | 11 (13%) / 36 (43.5%) / 35 (43.5%) |
| **cT category (%)** |  |
| cT1 / T2 / T3 | 1 (1%) / 3 (4%) / 78 (95%) |
| **cN category (%)** |  |
| cN0 / N1 / N2 | 30 (37%) / 32 (39%) / 20 (24%) |
| **cM category (%)** |  |
| cM0 / M1 | 79 (96%) / 3 (4%) |
| **cStage (%)** |  |
| I / II / III/ IV | 1 (1%) / 30 (37%) / 48 (58%) / 3 (4%) |
| **Recurrence (%)** |  |
| non-recurrence / recurrence | 37 (45%) / 45 (55%) |

### Target sequencing in clinical ESCC cases

A total of 56 autophagy- and ESCC-related genes were selected for original target enrichment sequencing. The autophagy-related genes were largely the same as those previously reported; however, some genes were added or deleted ^13,14^. The whole-genome analysis of 552 esophageal squamous cell carcinoma cases identified cancer driver genes. Among them, 19 genes with a high frequency of occurrence were selected ^15^. The target regions were designed to enrich the exonic regions and exon–intron junctions of all 56 genes (Table S1). The mean percentile of the covered target regions was 99.57%.

### DNA extraction, library preparation, and data analysis for targeted capture sequencing

Biopsy specimens from 82 patients were analyzed. The assessment and recovery of cancerous areas were performed using previously reported methods ^13,16^. From the extracted DNA, a library of all the exon sequences of the 56 genes was prepared using the HaloPlex Target Enrichment kit (Agilent Technologies, Santa Clara, CA, USA), according to the manufacturer’s instructions. The libraries were high-throughput sequenced on a NextSeq platform (Illumina, San Diego, CA, USA) with 150 bp paired-end reads according to the manufacturer’s protocol. The data were analyzed using previously reported methods ^13^. SNVs with multiple allelic characteristics were excluded. A violin plot was generated using the R package (https://bioconductor.org/packages/release/-bioc/html/edgeR.html).

### Study design and statistical analysis

A 2 × 2 cross-tabulation table was created with and without variants, and with and without recurrence. A Fisher’s exact test was performed using R based on the cross-tabulation table to examine the association between gene variants and recurrence.

The variants with a preliminarily inferred association with postoperative recurrence were examined for their associations with OS and recurrence-free survival (RFS). OS was defined as the period from the date of surgery to the date of death. RFS was defined as the period from the date of surgery to the date of first evidence of relapse. For patients who did not show relapse or die, RFS or OS was censored at the last confirmed date of no recurrence. OS and RFS were analyzed using the Kaplan–Meier method, and significance was determined by the log-rank test using the open-source Python software package. The median follow-up period for patients surviving without death was 51.2 (range: 7.0–107.8) months. Survival analysis was performed for 78 cases which had sufficient sample volume to allow the analysis of all variants that were candidates for prognostic factors. Univariate and Multivariate survival analyses were performed using a stratified Cox proportional hazard model. In the multivariate analyses, covariates were selected using backward elimination. All statistical tests were two-sided, and p < 0.05 was considered statistically significant.

### RNA extraction, library preparation, and data analysis for RNA sequence

Total RNA was isolated from formalin-fixed paraffin-embedded (FFPE) biopsy specimens (n = 15) from patients with esophageal cancer were treated between 2012 and 2019. Libraries for RNA sequencing were prepared from total RNA as described previously ^17^. Of the 15 specimens, eight and seven specimens showed non-recurrence and recurrence, respectively.

The resulting library was sequenced on an Illumina HiSeqX platform (2 × 150-bp read length). Data analysis was based on previously reported methods with some modifications ^17^. Differentially Expressed Genes (DEGs) were defined as genes that showed a two-fold or greater difference in the expression level of transcripts per million (TPM) values between the recurrence and non-recurrence groups and a significant difference of p < 0.05. The significance estimate of the differences in gene expression, such as the p-value, was calculated using expected counts from the RSEM software package using edgeR package in R, and a volcano plot figure was generated using R. The DAVID database (https://david.ncifcrf.gov/) was used for Gene Ontology (GO) analyses. STRING (https://stringdb.org/cgi/input?sessionId=b89PQA39oVO5&input_page_show_search=on) analysis was used to identify protein-protein interaction networks associated with highly expressed transcripts. Raw counts from the gene expression data were normalized to log counts per million (log-CPM) and further transformed into z-scores. Principal component analysis (PCA) and heat map plots were created using R, based on log-CPM (z-score) values.

### Machine learning model construction

The model development was performed using the Google Collab platform, and Pycaret was the first package used for machine learning, which required the installation of packages containing pandas, NumPy, warnings, and Pycaret (Moez A. PyCaret: an open-source, low-code machine learning library in Python. https://www.pycaret.org). Feature sets from eight different patient backgrounds and 21 different patient backgrounds + <u>SNVs were separately entered into Pycaret</u>. Pycaret divided each set into training (70%) and independent test cohorts (30%) to build a recurrence prediction model. Each feature set was trained on 15 machine learning models, and the stability of the models was evaluated by performing 10-fold cross-validation of the performance of each model and the genomic features that contributed the most to automatic generation in the training cohort. The most accurate models were subjected to hyperparameter tuning, and the tuned models were assembled using the blending method. The missing values in the SNVs were filled using Pandas df. fillna (data.mean()).

## Results

### Original target enrichment sequencing

Biopsy specimens from patients with esophageal cancer undergoing radical surgery after treatment with the CF regimen were used to identify SNVs and INDELs for 56 genes related to autophagy and esophageal cancer using targeted enrichment sequencing to construct a prognostic system for the CF regimen. Between May 2012 and June 2020, 91 patients underwent the CF regimen + radical surgery at Saitama Medical University International Medical Center, of which 82 patients were eligible for the study after the required amount of DNA was obtained. The clinical characteristics of the 82 patients are presented in Table 1. Among the 82 patients, 45 experienced recurrence, representing a 55% recurrence rate. Next-generation sequencing yielded a median of 2,252,009 reads per sample (range: 714,042–6,247,424 reads per sample). Among the designed target bases, 87.1% (range: 40.2–98.4% per sample) had at least a 15-fold coverage, with a mean coverage of 660 fold (range: 156.11–1,963 fold) per nucleotide in the coding region of the target gene (Fig 1A-B).

**Fig 1.**
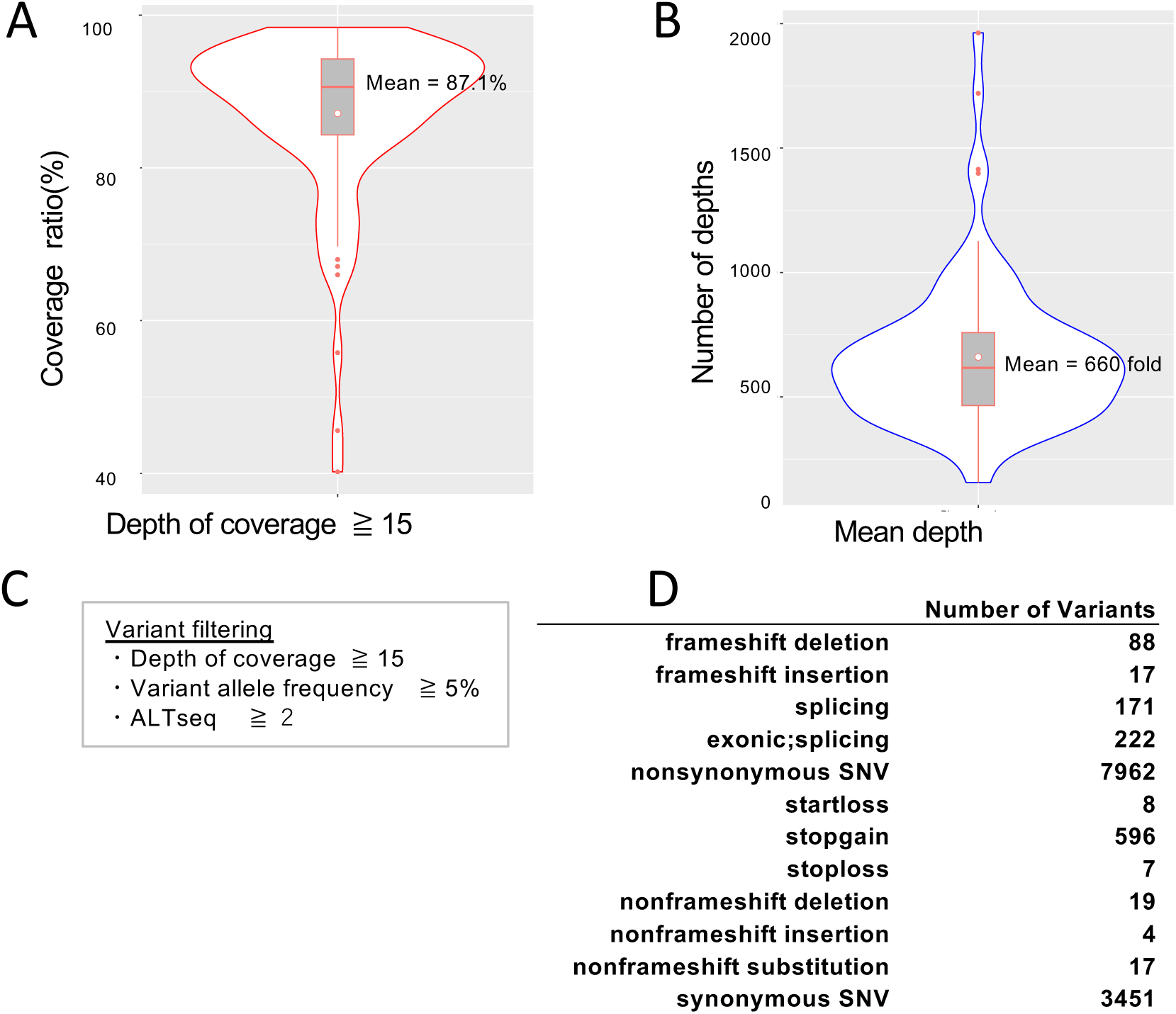
Results of the original target enrichment sequencing in our ESCC clinical cases. (A) The violin plot depicts the distribution of the coverage ratio for each of the 82 multiplexed samples. Percentage of regions had a depth of coverage greater than 15x . (B) The violin plot depicts the distribution of the mean depth for each of the 82 multiplexed samples. (C) Variant filtering thresholds (AltSeq: Alt, any other allele found at that locus). (D) The number of SNVs or INDELs identified by the original target enrichment sequencing is shown. The classification was performed by variant type.

### Breakdown of SNVs and INDELs

The next-generation sequencing data analysis only reported the presence of AltSeq (Alt, any other allele found at that locus); therefore, if there were no sequence reads, AltSeq was considered absent. Variant filtering based on criteria such as depth of coverage, variant allele frequency, and AltSeq counts reduces false-positive results and ensures confidence in the detected variants. By setting thresholds for "Depth of coverage ≥ 15, Variant allele frequency ≥ 5%, and AltSeq ≥ 2", we aimed to ensure that detected variants were supported by enough sequencing reads and were present at a significant level to be considered genuine (Fig 1C). The original target enrichment sequencing for cases with neoadjuvant chemotherapy showed that a total of 12,562 SNVs or INDELs were detected within the target region (Fig 1D). Among these variants, 7,962 were non-synonymous SNVs, indicating that they resulted in amino acid changes in the protein sequences. Additionally, 88 frameshift deletions and 17 frameshift insertions were detected, indicating that these alterations caused a shift in the reading frame of the gene. The SNVs associated with stop-gain variants were identified at 596 locations (Fig 1D). These variants resulted in premature termination of protein synthesis.

### Variants correlated with recurrence

We examined the associations among SNVs, INDELs, and recurrence. The variants found in a sample of 82 patients with recurrence were treated as binary events and subjected to Fisher exact tests. Thirteen variants were significantly different (p < 0.05). Among these variants, eight were non-synonymous SNVs, four were synonymous SNVs, and one was a splicing-site variant (Table 2).

**Table 2.** Results of target enrichment sequencing.

| Gene symbol | Exonic function | Nucleotide change | Aa change | <u>Non-recurrence</u> |  | <u>Recurrence</u> |  | p-value |
| --- | --- | --- | --- | --- | --- | --- | --- | --- |
|  |  |  |  | RefSeq (n) | AltSeq (n) | RefSeq (n) | AltSeq (n) |  |
| *EP300* | non-synonymous SNV | NM_001362843:c.5432G>A | p.R1811Q | 31 | 7 | 44 | 0 | 0.0072 |
| *PTCH1* | non-synonymous SNV | NM_001354918:c.4169G>A | p.R1390Q | 29 | 6 | 42 | 0 | 0.0062 |
| *ATG2A* | non-synonymous SNV | NM_001367971:c.1432C>T | p.R478C | 37 | 0 | 37 | 7 | 0.0147 |
| *ATG7* | synonymous SNV | NM_001144912:c.1857T>C | p.D619D | 33 | 5 | 45 | 0 | 0.0160 |
| *ULK2* | splicing | NM_001142610:c.1442-1G>T | splicing | 19 | 17 | 33 | 10 | 0.0306 |
| *BNIP3* | non-synonymous SNV | NM_004052:c.610C>T | p.R204C | 28 | 4 | 41 | 0 | 0.0330 |
| *FAT1* | non-synonymous SNV | NM_005245:c.6118G>A | p.D2040N | 32 | 4 | 44 | 0 | 0.0348 |
| *ULK2* | non-synonymous SNV | NM_001142610:c.1464C>A | p.F488L | 30 | 4 | 41 | 0 | 0.0356 |
| *ULK1* | non-synonymous SNV | NM_003565:c.2446A>G | p.T816A | 29 | 9 | 24 | 21 | 0.0416 |
| *BECN1* | synonymous SNV | NM_001313998:c.1302G>A | p.T434T | 34 | 4 | 45 | 0 | 0.0378 |
| *ZNF750* | synonymous SNV | NM_024702:c.618C>T | p.P206P | 34 | 4 | 45 | 0 | 0.0378 |
| *KMT2D* | non-synonymous SNV | NM_003482:c.12712C>T | p.R4238C | 34 | 4 | 44 | 0 | 0.0397 |
| *FAT1* | synonymous SNV | NM_005245:c.12900G>A | p.A4300A | 34 | 4 | 42 | 0 | 0.0440 |

Eighty-four patients were included in the analysis; Fisher's exact test of 560 SNVs or INDELs showed 5 SNVs with $p < 0.05$ .
Aa change: amino acid change, RefSeq: allele in the reference genome, AltSeq: Alt, any other allele found at that locus.

### Survival analysis of 13 candidate SNVs in RFS and OS

The RFS was analyzed for 13 identified variants. The results indicated significant differences in RFS for six of these variants (Fig 2A-B and Table S2). The OS analysis was also performed for the 13 identified variants, with significant differences observed for two variants, ATG2A p.R478C (p < 0.005) and ULK2 splice sites (p=0.05) (Fig 2C-D).

**Fig 2.**
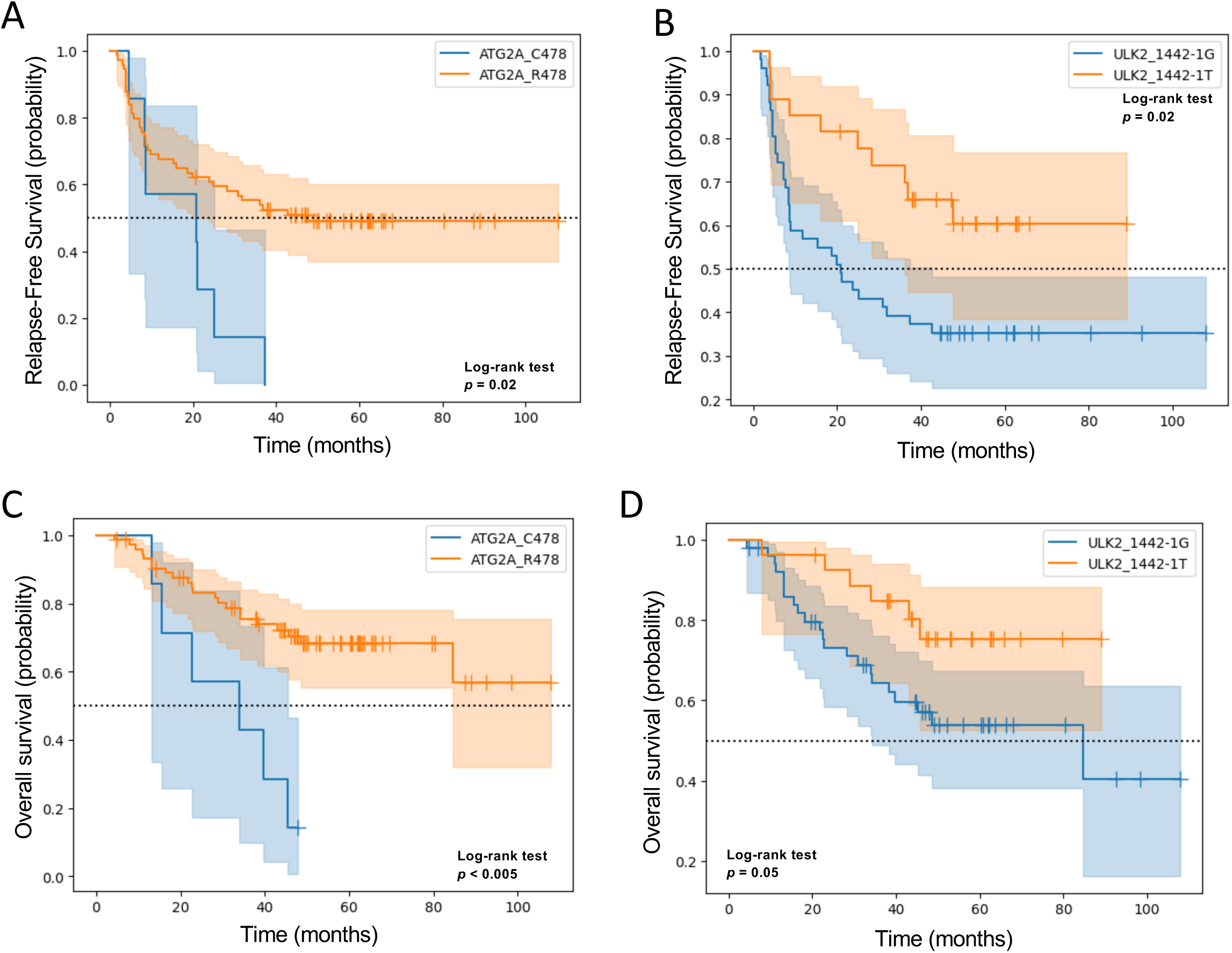
Relationship between variants of ATG2A p.R478C and ULK2 splice-site and ESCC prognosis with CF neoadjuvant chemotherapy. Relapse-free survival with (A) ATG2A p.R478C or (B) ULK2 1442-1 G> T, respectively. Overall survival with or without (C) ATG2A p.R478C or (D) ULK2 1442-1 G> T, respectively.

The variants in ATG2A p.R478C, extracted as candidate prognostic factors, showed an association with RFS and OS in univariate analysis (p=0.025 and 0.002, respectively). The variants in the ULK2 splice site were associated with RFS but not with OS (p=0.016 and 0.054, respectively). Additionally, multivariate Cox regression analysis revealed that the presence of variants in ATG2A_R478C or the absence of variants in ULK2_1442-2G>T (p=0.046, hazard ratio=2.076) and conventional open thoracotomy (p=0.018, hazard ratio=2.096) were independent prognostic factors for RFS. Likewise, multivariate Cox regression analysis of OS revealed that the presence of variants in ATG2A_R478C or the absence of variants in ULK2_1442-2G>T (p=0.029, hazard ratio=2.764) and clinical lymph node metastasis (p=0.040, hazard ratio=2.604) were independent prognostic factors for OS (Tables 3–4).

**Table 3.**
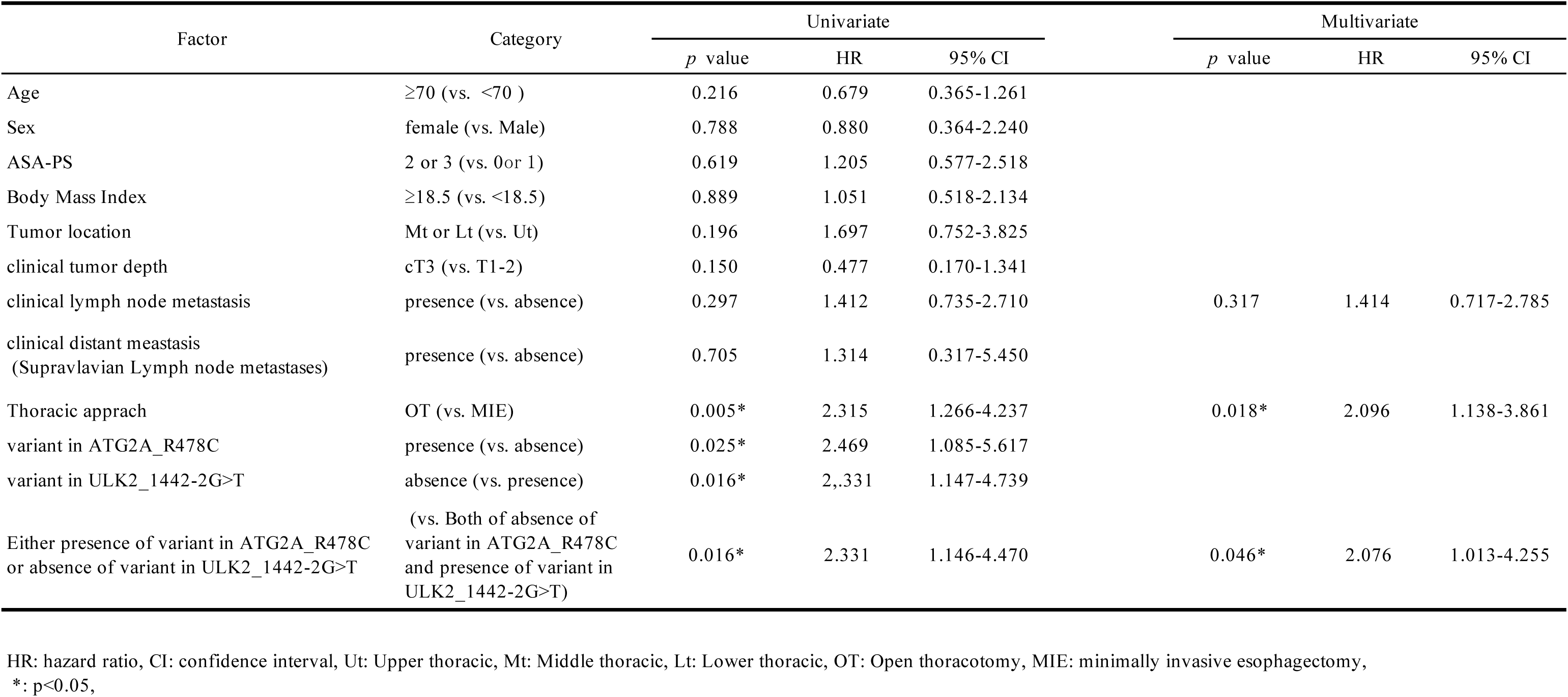
Univariate and multivariate Cox regression analysis for RFS.

**Table 4.** Univariate and multivariate Cox regression analysis for OS.

| Factor | Category | Univariate |  |  | Multivariate |  |  |
| --- | --- | --- | --- | --- | --- | --- | --- |
|  |  | *p* value | HR | 95% CI | *p* value | HR | 95% CI |
| Age | ≥70 (vs. <70 ) | 0.394 | 0.716 | 0.330-1.551 |  |  |  |
| Sex | female (vs. Male) | 0.834 | 0.880 | 0.264-2.927 |  |  |  |
| ASA-PS | 2 or 3 (vs. 0 or 1) | 0.302 | 1.651 | 0.636-4.284 |  |  |  |
| Body Mass Index | ≥18.5 (vs. <18.5) | 0.721 | 0.856 | 0.364-2.014 |  |  |  |
| Tumor location | Mt or Lt (vs. Ut) | 0.694 | 1.237 | 0.428-3.579 |  |  |  |
| clinical tumor depth | cT3 (vs. T1-2) | 0.862 | 1.194 | 0.162-8.819 |  |  |  |
| clinical lymph node metastasis | presence (vs. absence) | 0.077 | 2.210 | 0.896-5.451 | 0.040* | 2.604 | 1.047-6.476 |
| clinical distant metastasis<br>(Supravlavian Lymph node metastases) | presence (vs. absence) | 0.651 | 1.396 | 0.326-5.969 |  |  |  |
| Thoracic approach | OT (vs. MIE) | 0.050 | 2.119 | 0.983-4.566 | 0.212 | 1.650 | 0.751-3.623 |
| variant in ATG2A_R478C | presence (vs. absence) | 0.002* | 3.741 | 1.494-9.366 |  |  |  |
| variant in ULK2_1442-2G>T | absence (vs. presence) | 0.054 | 2.370 | 0.959-5.848 |  |  |  |
| Either presence of variant in ATG2A_R478C<br>or absence of variant in ULK2_1442-2G>T | (vs. Both of absence of<br>variant in ATG2A_R478C<br>and presence of variant in<br>ULK2_1442-2G>T) | 0.054 | 2.371 | 0.959-5.860 | 0.029* | 2.764 | 1.109-6.890 |

HR: hazard ratio, CI: confidence interval, Ut: Upper thoracic, Mt: Middle thoracic, Lt: Lower thoracic, OT: Open thoracotomy, MIE: minimally invasive esophagectomy,
\*. $p < 0.05$

### Correlation between pathogenic/likely pathogenic SNVs and recurrence rate

The SNVs and INDELs classified as pathogenic or likely pathogenic mutations in ClinVar were found at 212 locations in 22 genes, including 17 esophageal cancer-related genes. Among these variants, 11 were frameshift deletions, 15 were splicing variants, 95 were non-synonymous SNVs, and 87 were stop-gain variants. Fisher’s exact test was conducted for each gene to assess its association with recurrence; however, none of the genes showed statistically significant differences (Fig 1S). Moreover, pathogenic/likely pathogenic variants were observed in 81 of the 82 analyzed specimens. This suggests that while these variants were prevalent among the patient samples, they did not appear to significantly influence recurrence or prognosis.

### Machine learning model to predict recurrence

In this study, 13 SNVs were identified as candidate predictors of recurrence after neoadjuvant chemotherapy with the CF regimen for esophageal cancer. However, some specimens had multiple types of SNVs, thus, the question remained as to which SNVs should be trusted for prediction (Fig 2SA). Therefore, we investigated the construction of a recurrence prediction model using machine learning, considering 21 factors, including the SNVs found in this study and patient background (Fig 2SB).

Fifteen algorithms were trained using the Pycaret classification module, and 21 features, including patient background and SNV, to construct a model with recurrence as the correct answer. The accuracy level of the entire model was compared based on the accuracy value, and the results showed the highest value of 0.8467 for Naive Bayes (Table S3). Furthermore, when tune_model was used for accuracy, the value became 0.88, which was defined as the final_model (Fig 3A). When eight types of patient backgrounds were used as features, the accuracy was 0.5633 in Naïve Bayes (Fig 3A). Additionally, other evaluation metrics such as Recall, Precision (Prec.), F1 Score, Kappa, and Matthews Correlation Coefficient (MCC) also demonstrated the superiority of the model using all 21 factors, including SNVs (Fig 3A). These results showed that incorporating SNVs along with patient background information significantly improved the predictive performance of the model.

**Fig 3.**
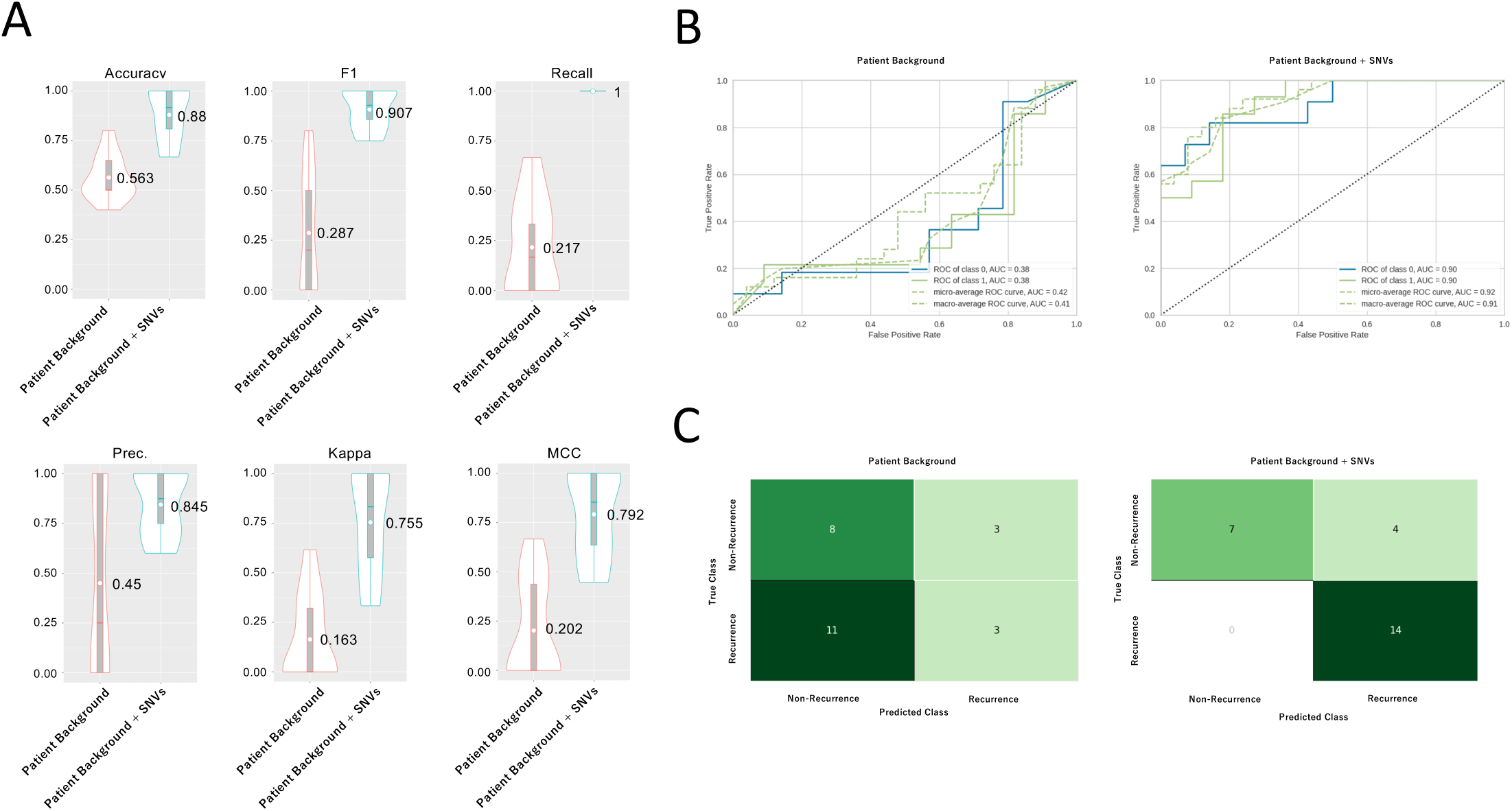
Machine learning model to predict recurrence. (A) An indicator to evaluate the prediction of recurrence by Naive Bayes with patient background or patient background + SNVs (single nucleotide variants) as features. (Prec.: Precision). (B) The ROC (receiver operating characteristic) curve for fine-tuned Naive Bayes. Class 0 implies non-recurrence. Class 1 implies recurrence. (C) Confusion matrix for fine-tuned Naive Bayes.

A Receiver Operating Characteristic (ROC) curve for Naive Bayes was generated, showing an Area Under the Curve (AUC) value of 0.9 for both class 0 (no recurrence) and class 1 (recurrence) (Fig 3B). This AUC value indicated that this model had an excellent discriminative ability to distinguish between possible recurrences.

The confusion matrix was one of the representations used in machine learning to evaluate the performance of classification models. From the confusion table, the true positive value was seven, which was higher than the false negative value of four (Fig 3C). Furthermore, it is noteworthy that the number of true negatives was 14 and the number of false positives was 0 (Fig 3C). These results suggested that the Naive Bayes classification model had high performance in terms of both sensitivity (true positive rate) and specificity (true negative rate) in predicting recurrence in patients with esophageal cancer.

### Comparison of the expressions of coding RNAs between recurrence and non-recurrence groups

This study demonstrated several predictive systems for ineffective CF regimens. Therefore, to propose a selective treatment for the poor response group, we performed a comprehensive expression analysis to understand the biological characteristics of the poor response group. In the analysis of differential gene expression between recurrence and non-recurrence specimens among 19,972 coding genes, 187 genes were found to have higher expression levels in the recurrence group compared to the non-recurrence group, whereas 128 genes showed lower expression levels in the recurrence group compared to the non-recurrence group based on the criteria of fold change ≥ 2 and p-value < 0.05 (Fig 4A).

**Fig 4.**
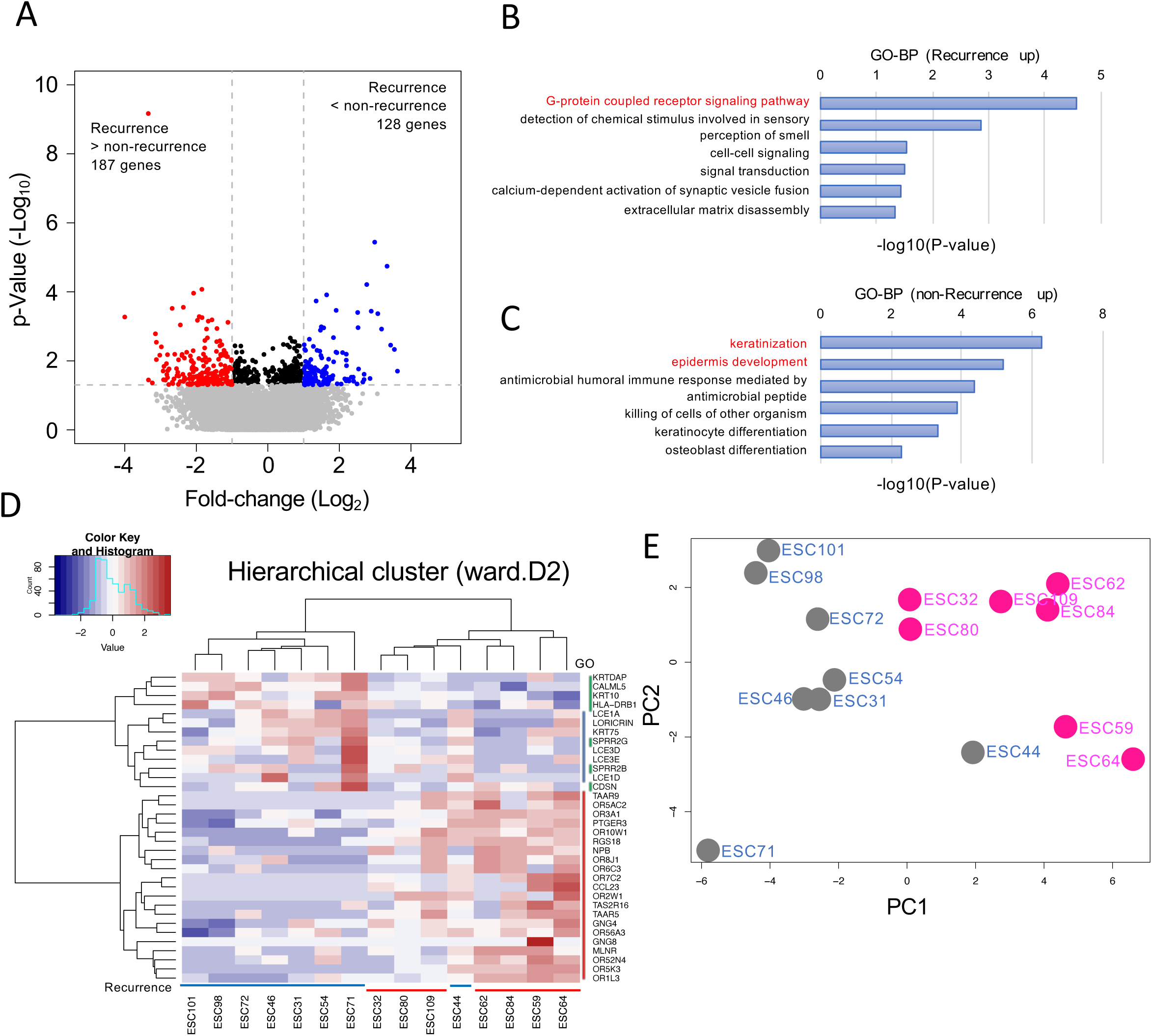
Expression analysis between recurrence and non-recurrence groups. (A) Volcano plot of differentially expressed genes between recurrence (n=7) and non-recurrence (n=8) groups. Summary of biological processes in gene ontology analysis (GO-BP) of genes with elevated expression in recurrence (B) and non-recurrence (C) groups. (D) Heatmap and hierarchical cluster analysis performed on 34 genes. Blue above specimen number means no recurrence, red means recurrence. The color coding next to the gene means that the gene is included in the G-protein coupled receptor signaling pathway (red), keratinization (blue), and epidermis development (yellow). (E) Principal component analysis performed on 34 genes. Red specimen name indicates recurrence group, gray specimen name indicates non-recurrence group.

GO analysis using the DAVID database revealed the enrichment of specific biological processes associated with highly expressed genes in the recurrence and recurrence-free groups. In the recurrence group, 21 genes with high expression were enriched in processes related to the "G-protein coupled receptor (GPCR) signaling pathway" (Fig 4B). In contrast, in the recurrence-free group, eight and seven genes with high expression were enriched in processes related to "keratinization" and "epidermis development," respectively (Fig 4C). These genes, especially those contributing to the "GPCR signaling pathway," may be potential additional therapeutic targets for patients with poor response to the CF regimen.

### Possibility of predicting recurrence by analysis of gene set expression levels

Recurrence was predicted by analyzing the expression levels of 34 genes associated with the three GO terms. Initially, the expression levels of the 34 genes among the samples were visualized using a heat map to gain insight into the differences and similarities between the samples. Hierarchical cluster analysis was used to group the samples based on similarities in their gene expression profiles. The results showed that except for the 44 ESC samples, the samples predominantly clustered into two main groups: recurrence and non-recurrence (Fig 4D). Principal component analysis (PCA) was performed on the expression data of 34 genes. Plotting the data in the first two principal components showed that, excluding the 44 ESC samples as in the hierarchical cluster analysis, recurrence could effectively distinguish non-recurrence samples in the first principal component (Fig 4E).

The expression levels of 56 genes related to autophagy and esophageal cancer were also examined. Only CDKN2A showed a significant difference at p > 0.02, with an approximately 7.6-fold increase in expression in the recurrent group compared to that in the non-recurrence group (Fig 4A). Clustering and PCA were performed on 37 autophagy-related genes (Fig 3S A-B); however, the results did not clearly distinguish non-recurrence from relapse. This suggested that the 56 genes associated with autophagy and esophageal cancer were not compatible with the prediction of recurrence by expression level analysis.

## Discussion

Although neoadjuvant chemotherapy with DCF therapy, a three-drug combination regimen for resectable locally advanced esophageal cancer, is expected to prolong the prognosis of postoperative survival, the high incidence of adverse events, such as myelosuppression, is a major clinical issue. CF, a conventional neoadjuvant chemotherapy regimen, showed superior tolerability in terms of drug toxicity, although its prolonged postoperative prognostic effect was less satisfactory than that of DCF therapy. We identified various autophagy- and esophageal cancer-related gene variants as biomarkers that could predict the efficacy of CF therapy prior to treatment. Eventually, we constructed a machine learning model that was highly predictive of postoperative recurrence based on 21 factors consisting of clinical factors and SNVs. We also established a highly heterogeneous treatment selection system using the machine-learning model.

In surgical specimens from patients with esophageal cancer receiving neoadjuvant chemotherapy, a high expression of PINK1 protein, an initiator of mitophagy, correlated with poor response to neoadjuvant chemotherapy with CF or DCF regimen; however, this correlation was not observed in biopsy specimens taken prior to chemotherapy. Thus, PINK1 protein expression is not considered a predictive biomarker for response to neoadjuvant chemotherapy with CF or DCF regimen in patients with esophageal cancer. In contrast to PINK1 protein expression, the 13 SNVs identified in this study were prognostic predictors of neoadjuvant chemotherapy with the CF regimen in patients with esophageal cancer. Specifically, SNVs such as p.R478C in ATG2A and in the splice site of ULK2 were found to be significant. These SNVs are reported for the first time as prognostic predictors of esophageal cancer. However, SNVs in PINK1 (c.1018G>A and c.1562A>C), which were previously suggested to be prognostic factors for 5-FU-based adjuvant chemotherapy in colon cancer, showed no significance in ESCC. Conversely, SNVs identified in esophageal cancer did not show significant differences in colorectal cancer. The difference in the prognosis-related SNVs between colorectal cancer and ESCC suggests that the genetic characteristics affecting treatment response and outcome may differ significantly between different cancer types. This underscores the importance of considering organ-specific genetic profiles when developing personalized medical approaches.

ATG2A plays an important role in autophagosome formation, an early step in autophagy, and promotes the lipid translocation required for autophagosome membrane expansion^18^. ATG2A promotes colony formation and migration in glioblastoma cell lines by activating autophagy. This suggests that it is involved in cancer progression and therapeutic response^19^. ATG2A p.C478 minor variant was found to be significantly associated with worse RFS and OS compared to p.R478 major variant. This suggests that the p.C478 variant may contribute to a poor response to CF regimens by activating autophagy. In silico analysis with PolyPhen2, the p.C478 variant was "probably damaging," with a score of 1.000, indicating that the high score was functionally significant. Further biochemical characterization is needed to better understand the functional impact and role of p.C478 mutation in ATG2A during neoadjuvant chemotherapy for esophageal cancer. Such characterization efforts may pave the way for the development of targeted therapies aimed at modulating the activity of this mutant and for the identification of biomarkers to guide treatment decisions, especially in relation to CF regimens. Thirteen variants showed significant differences in predicting recurrence after treatment with CF regimens for esophageal cancer. Therefore, we constructed a machine learning method to predict recurrence using 21 features, including eight patient backgrounds and 13 SNVs, which showed high accuracy (0.88) and AUC (0.9).

However, this study had some limitations. First, the differences in the importance of the features of the 13 SNVs were examined; however, the Naïve Bayes algorithm was difficult to compute directly and has not been shown. Second, in this study, the hyperparameters were optimized collectively using the tune_model function; however, it was also possible to effectively tune the individual parameters. Third, the number of samples was limited because the test sample was 25, and the cohort was single. In contrast, PCA and hierarchical clustering analyses of 34 genes identified as DEGs in the expression analysis of recurrent and non-recurrent groups suggested the possibility of predicting recurrence. In the confusion matrix shown in Figure 3C, there were four cases of non-recurrence in which recurrence was expected. In the future, we would like to consider improving the decision rate when SNVs analysis and pathological image results are added to machine learning features.

Prognostic prediction of CF regimens by SNVs showed that the ATG2A p.R478C (p < 0.005) and ULK2 splice site (p=0.05) variants were candidates. Furthermore, when machine learning was performed, considering information from the 13 SNVs as features, the AUC=0.9 was high. As the purpose of the RNA-seq analysis in this study was to determine the biological characteristics of the recurrent group, the number of samples was limited to 15. In the future, the number of specimens should be 82, the same as in the SNVs analysis, to examine whether prognosis can be predicted by expression analysis. We would also like to consider improving the decision rate when the results of the expression analysis are added to the machine learning features.

Genes contributing to the GPCR signaling pathway were enriched in the recurrent group. They are located on the cell membrane and transduce extracellular signals to produce key physiological effects^20^. Activated by external signals through coupling to different G proteins or arrestins, GPCRs elicit a cyclic adenosine 3,5-monophosphate (cAMP) response, calcium mobilization, and phosphorylation of extracellular signal-regulated protein kinases 1/2(pERK1/2)^21,22^. As one of the most successful therapeutic target families, GPCRs have undergone a transition from random ligand screening to knowledge-driven drug design^21^. Of the 826 human GPCRs, approximately 350 were regarded as druggable and 165 were validated drug targets^21^. GNG4, which was highly expressed in the recurrent group, is a member of the G-protein γ family, which typically transduces signals from upstream GPCRs^23^. GNG4 is upregulated in primary gastric cancer and liver metastatic lesions. High expression of GNG4 in primary cancer tissues are associated with shorter OS and likelihood of liver recurrence. Functional assays revealed that GNG4 promoted cancer cell proliferation, cell cycle, and adhesion^24^. Tumor formation in GNG4-knockout cells was moderately reduced in a subcutaneous mouse model and strikingly attenuated in a mouse model of liver metastasis^24^. Genes contributing to the GPCR signaling pathway that were highly expressed in the recurrent group, particularly GNG4, are potential drug targets.

The other limitations were that this study was a small-sample single-center study, and future validation with a larger cohort is needed. Second, the median follow-up period of surviving patients was relatively short, and further follow-up studies are required to evaluate the long-term results of this study. Third, the biochemical functions of the variants identified in this study are unknown.

Nonetheless, we believe that our model for predicting the efficacy of CF therapy is significant because it is expected to avoid excessive drug toxicity caused by DCF therapy while simultaneously providing a non-inferior therapeutic effect. The therapeutic effect of standard DCF therapy can be expected even in cases in which CF therapy is deemed ineffective. In future studies, it will be necessary to examine whether our model for predicting the efficacy of CF therapy is relevant to predict the efficacy of DCF therapy, particularly if patients who are expected to receive ineffective CF therapy can be rescued by DCF therapy.

In conclusion, candidate genes were identified to predict the prognosis of CF regimens, and a machine learning model was constructed to further predict recurrence. We believe that this information will be useful for the selection of neoadjuvant　chemotherapy, including the avoidance of docetaxel. The avoidance of unnecessary drugs may provide useful guidance not only for patients, but also for health economics.

## Data Availability

Availability of data and materials
The data are not publicly available because of privacy and ethical restrictions. Access to the data and calculation methods can be obtained from the corresponding author upon request via email.

## Acknowledgments

We thank the staff of the Division of Analytical Science, Hidaka Branch of the Biomedical Research Center, Saitama Medical University, for providing research equipment and offering important advice. We would like to thank Editage (www.editage.jp) for English language editing.

## Availability of data and materials

The data are not publicly available because of privacy and ethical restrictions. Access to the data and calculation methods can be obtained from the corresponding author upon request via email.

## Disclosure

### Ethics Statement

- **Approval of the research protocol by an Institutional Reviewer Board.** This study adhered to the ethical standards of the Declaration of Helsinki and its subsequent amendments. This study was approved by the Institutional Review Board of the Saitama Medical University International Medical Center (2022-113 and 2024-055).
- **Informed Consent.** The requirement for informed consent was waived by the Institutional Review Board of Saitama Medical University International Medical Center in view of the retrospective nature of this study.
- **Registry and the Registration No. of the study/trial.** *N/A.*
- **Animal Studies.** *N/A*.

## Author Contributions

YM collected patient data, performed statistical analysis, and prepared the manuscript. MH performed the chromosomal DNA and total RNA extraction, analyzed and interpreted the data from the next-generation sequencer, and prepared the manuscript. YK prepared sections of FFPE samples. TK determined the tumor area. YB is an advisor for machine learning. TS and YS are advisors for bioinformatics. HF, KU, YM, HS and TH designed and supervised the study and revised the manuscript. All the authors have read and approved the final version of the manuscript.

## Abbreviations

CF: cisplatin + 5-fluorouracil
DCF: docetaxel + CF
OS: overall survival
RNA-seq: RNA sequencing
SNV: single nucleotide variant
AUC: area under the curve
ROC: Receiver Operating Characteristic
CIL: chemotherapy-induced leukopenia
RFS: recurrence-free survival
INDELs: insertions and deletions
ESCC: esophageal squamous cell carcinoma

## Supporting information

**Fig. S1.**
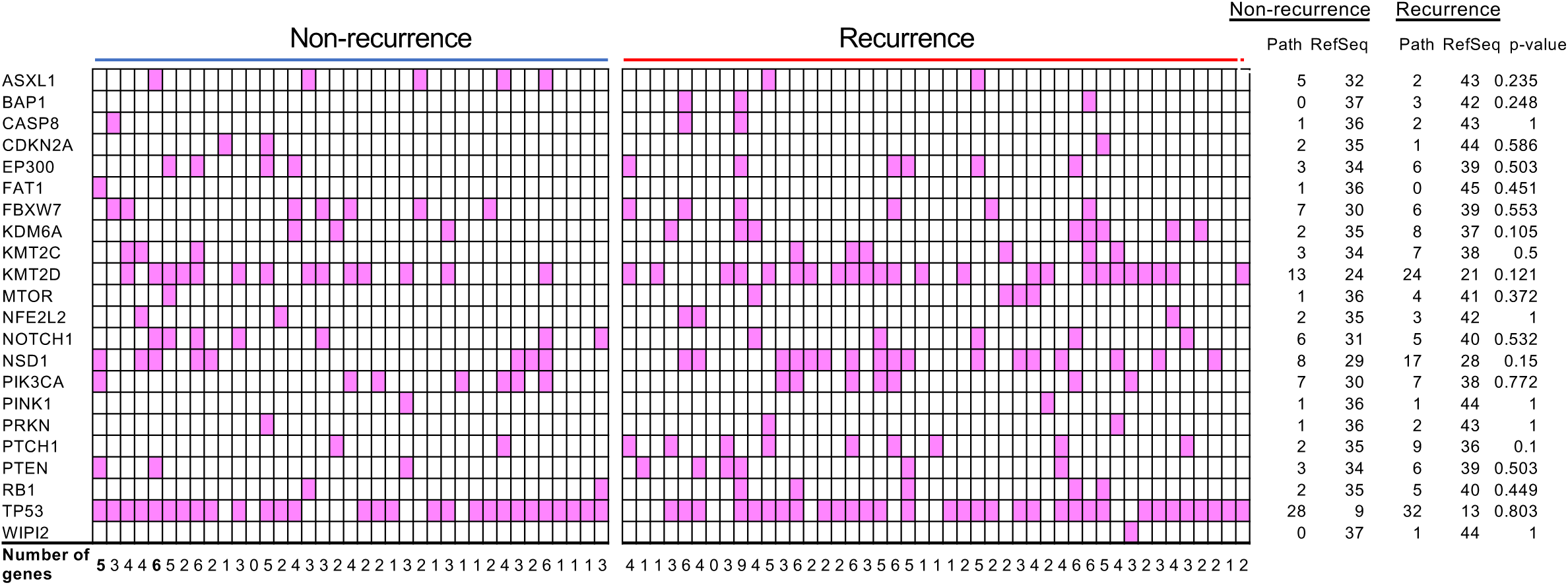
ClinVar-based pathogenic SNVs. Samples with variants defined as pathogenic/likely pathogenic by ClinVar are shown in red. Fisher’s exact test was performed for each gene that was determined to be pathogenic/likely pathogenic by ClinVar. Path: Pathogenic/Likely Pathogenic variant. RefSeq: allele in the reference genome.

**Fig. S2.**
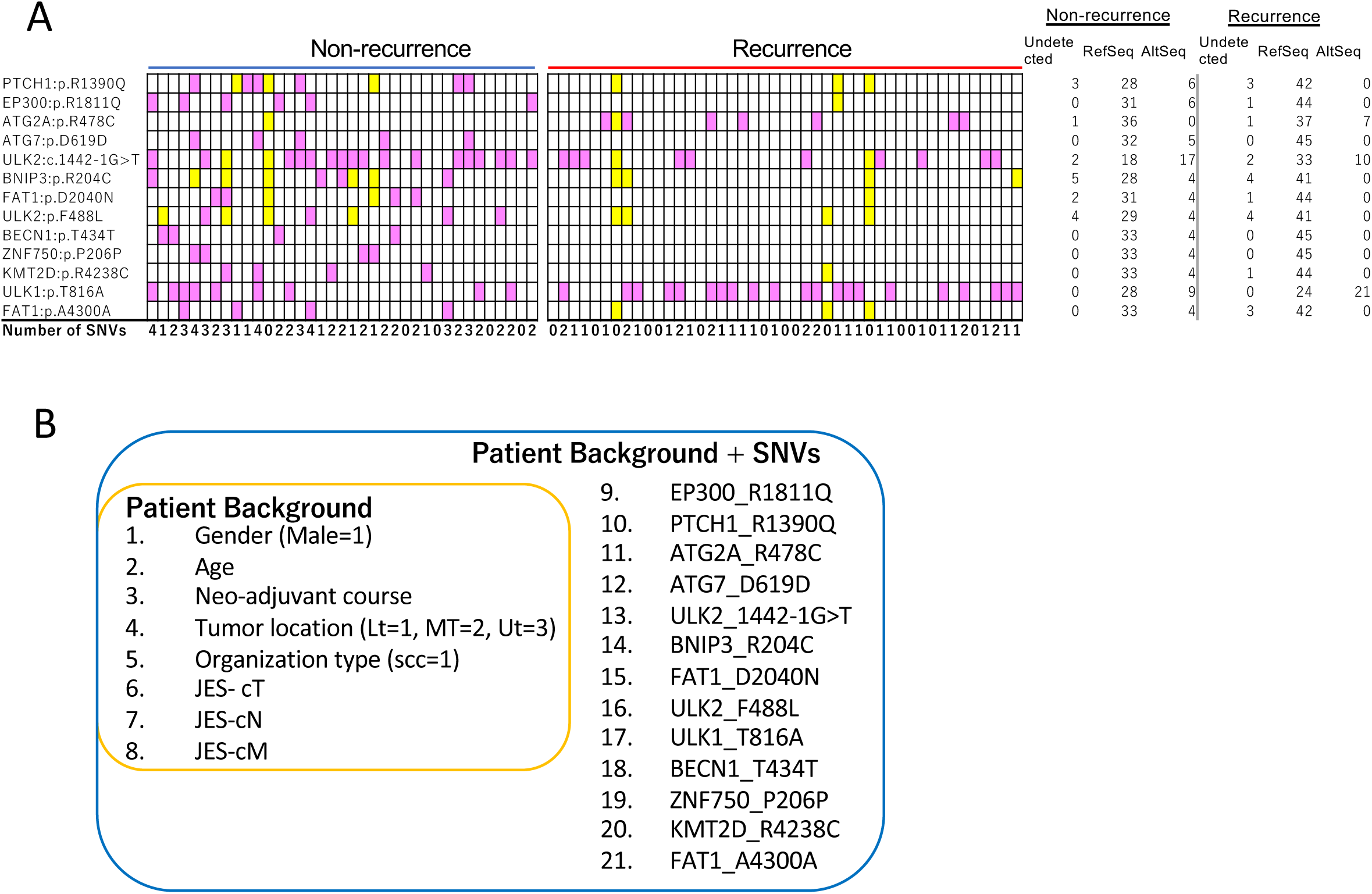
Variant distribution of specimens and elements of machine learning. (A) Samples with AltSeq variants are shown in red. Undetected samples with sequence reads that do not meet the criteria are shown in yellow. (B) Patient background only is circled in yellow. Patient background with additional SNVs information is circled in blue.

**Fig. S3.**
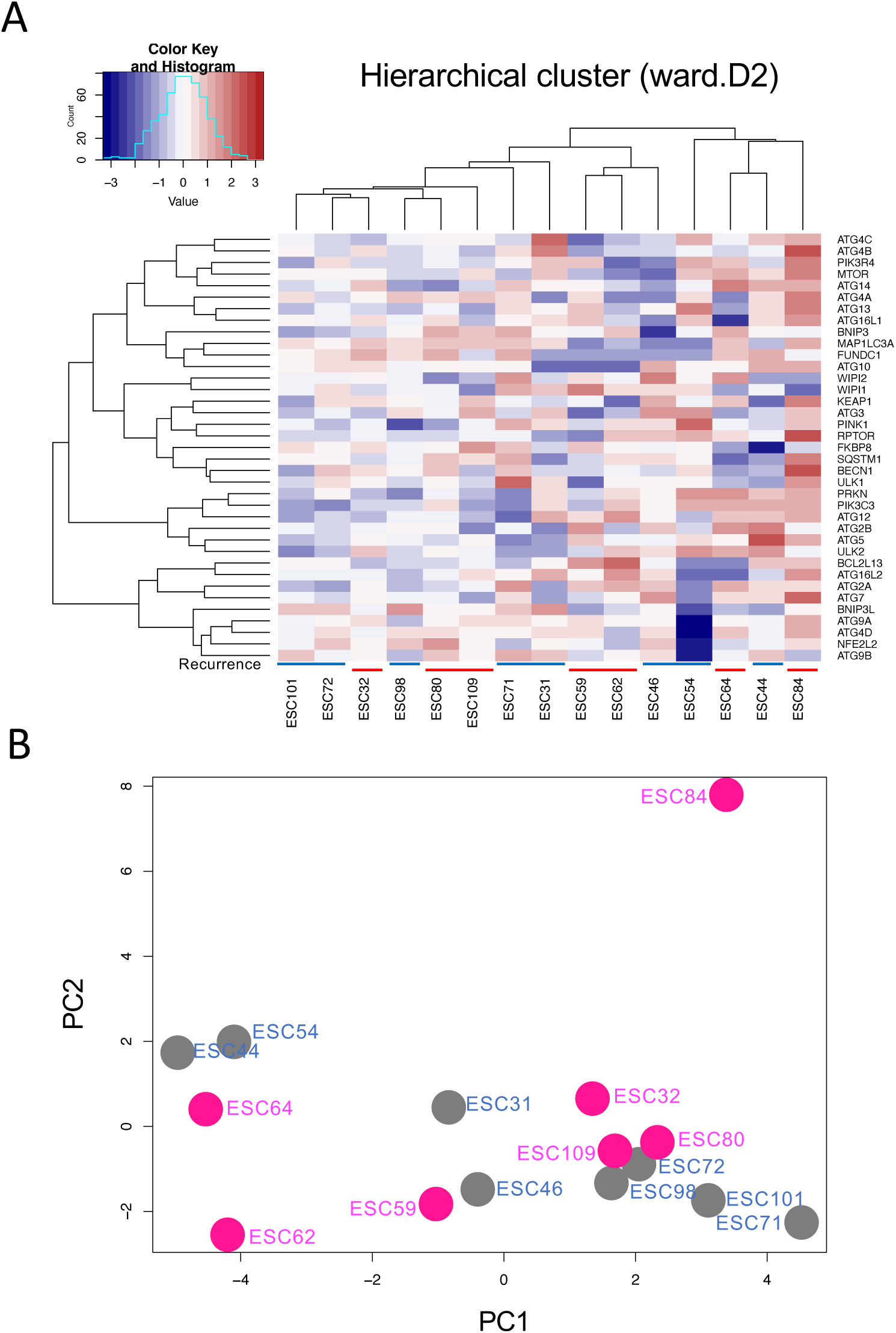
Expression analysis of autophagy-related genes. (A) Heatmap and hierarchical cluster analysis performed on 37 autophagy-related genes. Blue above specimen number indicates no-recurrence, red indicates recurrence. (B) Principal component analysis performed on 37 genes. Red specimen name indicates recurrence group, gray specimen name indicates no-recurrence group.

**Table S1.**
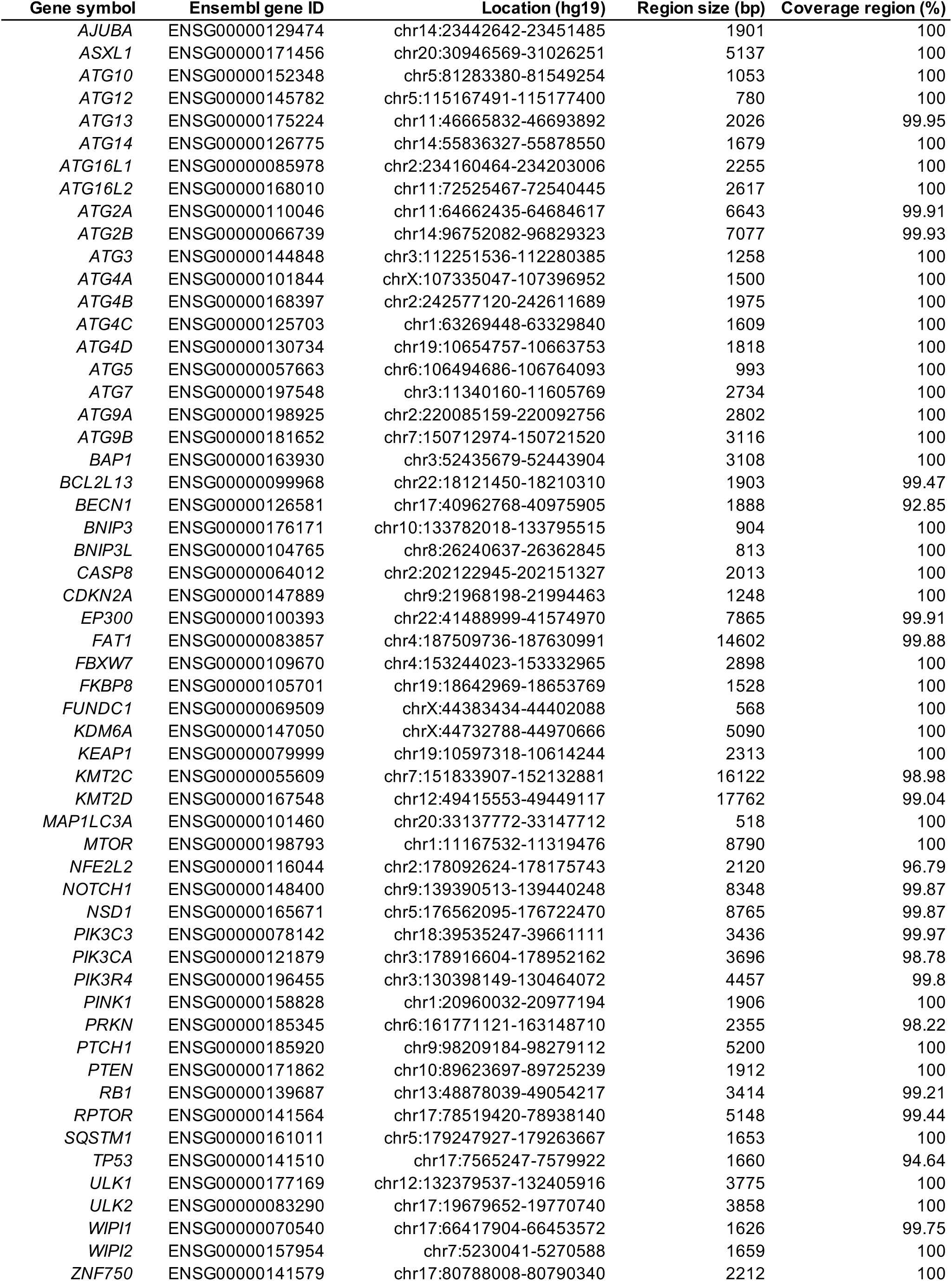
Targeted genes in clinical cases of esophageal cancer.

**Table S2.** Overall survival (OS) and recurrence-free survival (RFS) analysis of 13 identified variants.

| Gene symbol | Nucleotide change | Aa change | OS | RFS |
| --- | --- | --- | --- | --- |
| *EP300* | NM_001362843:c.5432G>A | p.R1811Q | 0.06 | **0.02** |
| *PTCH1* | NM_001354918:c.4169G>A | p.R1390Q | 0.1 | **0.02** |
| *ATG2A* | NM_001367971:c.1432C>T | p.R478C | **<0.005** | **0.02** |
| *ATG7* | NM_001144912:c.1857T>C | p.D619D | 0.14 | **0.04** |
| *ULK2* | NM_001142610:c.1442-1G>T | splicing | **0.05** | **0.02** |
| *BNIP3* | NM_004052:c.610C>T | p.R204C | 0.18 | 0.07 |
| *FAT1* | NM_005245:c.6118G>A | p.D2040N | 0.14 | 0.06 |
| *ULK2* | NM_001142610:c.1464C>A | p.F488L | 0.16 | 0.06 |
| *ULK1* | NM_003565:c.2446A>G | p.T816A | 0.08 | 0.06 |
| *BECN1* | NM_001313998:c.1302G>A | p.T434T | 0.14 | 0.06 |
| *ZNF750* | NM_024702:c.618C>T | p.P206P | 0.16 | 0.06 |
| *KMT2D* | NM_003482:c.12712C>T | p.R4238C | 0.15 | 0.06 |
| *FAT1* | NM_005245:c.12900G>A | p.A4300A | 0.15 | 0.07 |

**Table S3.**
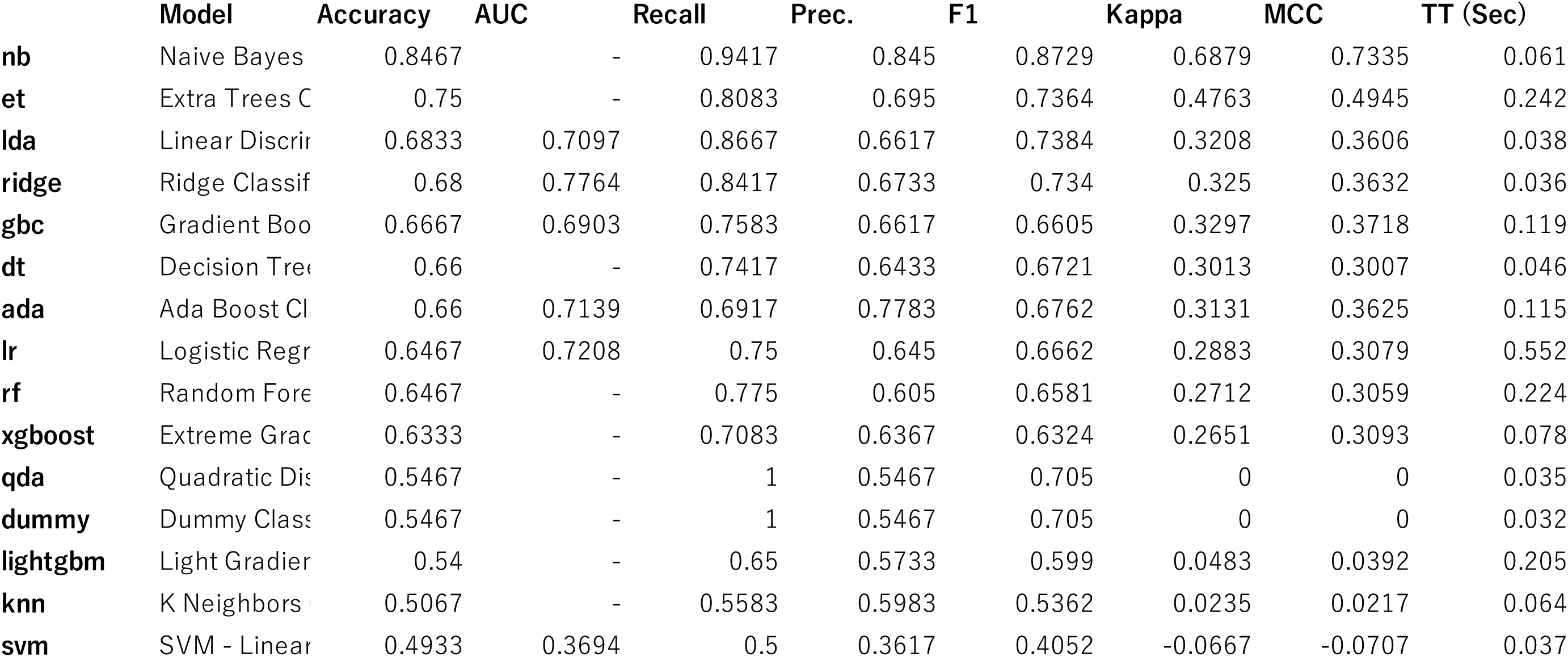
Results of the Pycaret classification module when Patient Background + SNVs is the feature and recurrence is the correct answer.

|  | **Model** | **Accuracy** | **AUC** | **Recall** | **Prec.** | **F1** | **Kappa** | **MCC** | **TT (Sec)** |
| --- | --- | --- | --- | --- | --- | --- | --- | --- | --- |
| **nb** | Naive Bayes | 0.8467 | - | 0.9417 | 0.845 | 0.8729 | 0.6879 | 0.7335 | 0.061 |
| **et** | Extra Trees C | 0.75 | - | 0.8083 | 0.695 | 0.7364 | 0.4763 | 0.4945 | 0.242 |
| **lda** | Linear Discrim | 0.6833 | 0.7097 | 0.8667 | 0.6617 | 0.7384 | 0.3208 | 0.3606 | 0.038 |
| **ridge** | Ridge Classif | 0.68 | 0.7764 | 0.8417 | 0.6733 | 0.734 | 0.325 | 0.3632 | 0.036 |
| **gbc** | Gradient Boo | 0.6667 | 0.6903 | 0.7583 | 0.6617 | 0.6605 | 0.3297 | 0.3718 | 0.119 |
| **dt** | Decision Tree | 0.66 | - | 0.7417 | 0.6433 | 0.6721 | 0.3013 | 0.3007 | 0.046 |
| **ada** | Ada Boost Cl | 0.66 | 0.7139 | 0.6917 | 0.7783 | 0.6762 | 0.3131 | 0.3625 | 0.115 |
| **lr** | Logistic Regr | 0.6467 | 0.7208 | 0.75 | 0.645 | 0.6662 | 0.2883 | 0.3079 | 0.552 |
| **rf** | Random Fore | 0.6467 | - | 0.775 | 0.605 | 0.6581 | 0.2712 | 0.3059 | 0.224 |
| **xgboost** | Extreme Grac | 0.6333 | - | 0.7083 | 0.6367 | 0.6324 | 0.2651 | 0.3093 | 0.078 |
| **qda** | Quadratic Dis | 0.5467 | - | 1 | 0.5467 | 0.705 | 0 | 0 | 0.035 |
| **dummy** | Dummy Class | 0.5467 | - | 1 | 0.5467 | 0.705 | 0 | 0 | 0.032 |
| **lightgbm** | Light Gradier | 0.54 | - | 0.65 | 0.5733 | 0.599 | 0.0483 | 0.0392 | 0.205 |
| **knn** | K Neighbors | 0.5067 | - | 0.5583 | 0.5983 | 0.5362 | 0.0235 | 0.0217 | 0.064 |
| **svm** | SVM - Linear | 0.4933 | 0.3694 | 0.5 | 0.3617 | 0.4052 | -0.0667 | -0.0707 | 0.037 |

